# Knowledge regarding safe abortion among married women of reproductive age group in Vyas Municipality 9, Tanahun

**DOI:** 10.64898/2026.08.26.26361402

**Authors:** Anju Bhujel, K.C Prisha, Dipesh Thapa

**Author notes:** Corresponding author: Dipesh Thapa. Main author: Anju Bhujel. Assistant Lecturer/ Research Guide: Prisha K.C.

## Abstract

**Background:** Abortion is safe when carried out using a method recommended by the World Health Organization (WHO), appropriate to the pregnancy duration, and by someone with the necessary skills. Globally, around 73 million induced abortions take place each year. Around 95% of maternal deaths occur in developing countries due to childbirth and pregnancy-related complications.

**Methodology:** A descriptive cross-sectional study was used for the study among the married reproductive (20-45 years) age women using a non-probability purposive sampling technique. A self-developed semi-structured questionnaire was used via face-to-face interview for data collection. The collected data were analyzed using SPSS 16.0 version descriptive and inferential statistics were used to find the association between variables.

**Results:** The findings of the study show that among 118 respondents, nearly two-thirds (57.6%) of the respondents had adequate knowledge and less than half (42.4%) of the respondents had inadequate knowledge regarding safe abortion. The study further shows that there was a significant association between the level of knowledge regarding safe abortion and education status and education level with p-value <0.05.

**Conclusion:** The study concludes that nearly two third of the respondents have adequate knowledge regarding safe abortion. Educational status significantly influenced their level of knowledge. Thus, we could provide correct knowledge, education, and enhance awareness to married women in the reproductive age group.

## Introduction

Abortion is a termination of pregnancy (spontaneous, therapeutic, or induced) before the stage of viability, usually before 22^nd^ weeks of gestation (fetal weight < 500g) (Dutta D. *et al.,* 2004). Abortion is safe when carried out using a method recommended by the World Health Organization (WHO), appropriate to the pregnancy duration, and by someone with the necessary skills. However, when women with unwanted pregnancies face barriers to obtaining quality abortion, they often resort to unsafe abortion (Abortion World Health Organisation; 2024). Globally, around 73 million induced abortions take place each year. Six out of 10 (61%) of all unintended pregnancies, and 3 out of 10 (29%) of all pregnancies, end in induced abortion (Abortion World Health Organisation; 2024). A large number of reproductive age women die due to childbirth and pregnancy related complications, and of them, around 95% of maternal death occur in developing countries (Naidoo M *et al*., 2017).

Abortion may be spontaneous or induced, unsafe induced abortion, in particular, can result in severe health complications, including hemorrhage, sepsis, and perforation of the uterus (Diedrich J *et al.,* 2009). Around 95% of maternal death occurs in developing countries due to childbirth and pregnancy-related complications (Naidoo M *et al.,* 2017). In Nepal, abortion is reported to be the third leading cause of maternal death (Yogi A *et al.,* 2018). Nepal reported high maternal mortality rates, that is 239 deaths per 100,000 live births, in 2016, which were linked to unsafe abortion practices, according to the Nepal Demographic and Health Survey (NDHS 2016) (Ghimire PR *et al.,* 2019). Although Nepal legalized abortion in 2002, a significant number of women continue to access unsafe abortions. Unsafe abortion continues to be a leading contributor to maternal mortality. Women in the reproductive age group are at significant health risk from unwanted pregnancies and unsafe abortion procedures (Right to Safe Abortion in Nepal: Gender and Health Hub; 2023).

Abortion can have serious consequences on reproductive health, placing women in life-threatening conditions and contributing to increased maternal mortality rates (Acharya A *et al.,* 2017). In every country in the world, women with unwanted pregnancies seek induced abortions, safe or otherwise. Unsafe methods of abortion, particularly invasive methods, can kill and injure women, and some 13% of all maternal deaths are the result of abortions with unsafe methods provided in sub optimal conditions by unskilled providers (Say L *et al.,* 2014). According to Health management and information system (HMIS) annual report Fiscal Year 2078/79 abortion rate is increased by 2,728 cases in FY 2079/80 (93,463) as compared to FY 2078/79 (90,733); from FY 2077/78 (79,952), the increment in FY 2078/79 was 10,781 cases (HMISreport et al.,). Safe abortion services help to reduce deaths and morbidity from complications such as sepsis and uterine perforation leading to hysterectomy, arising from unsafe invasive procedures, in places where most abortions are still illegal. Adequate level of knowledge and attitude of reproductive age women on medication abortion contributes to prevention of unsafe abortion (Warriner IK *et al.,* 2006).

## METHODOLOGY

### Research Design

A descriptive cross-sectional research design based on a quantitative approach was used.

### Research Setting

The research was conducted in Vyas Municipality-09, Tanahun.

### Study population

The population for the study was married women of reproductive age group of Vyas Municipality-09, Tanahun.

### Sample Size

Sample size was estimated based on a cross-sectional study which was conducted among 255 reproductive age women in a slum area of Kathmandu, which found 92.5% had a good knowledge on safe abortion (Manandhar N *et al.,* 2021). All the data were collected using a questionnaire. The questionnaire comprised 26 questions, and participants were asked to self-complete the survey. Over all questions were written in English and Nepali, for the easy understanding and participation was anonymous and voluntary (Thapa, D *et al.,* 2026).

#### Sample size

According to Cochran’s formulae, n= z^2^pq/L^2^ (Manandhar N *et al.,* 2021)

Where,

n = sample size needed
Z= critical value for 95% confidence limit i.e. 1.96 P= prevalence rate is 92.5% (0.925)
q= 1-p

Now, calculating sample size for descriptive cross-sectional study in infinite population;

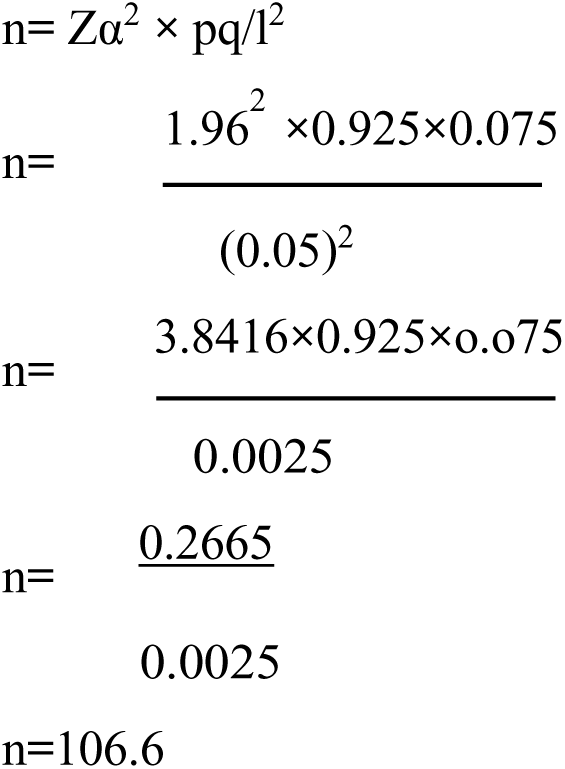

Therefore, the sample size (n) = 107

Taking 10% non-respondent rate

Final sample size= 107+ (10% of 107) = 118

### Sampling Techniques

Non-probability purposive sampling technique was used for the study.

### Inclusion Criteria

Those who were available to participate during data collection. Those who were willing to participate in the study.

### Exclusion Criteria

Those who were unable to provide information due to physical or mental illness.

### Tool of Data Collection

A self-developed semi-structured questionnaire was used to assess the knowledge regarding safe abortion. The research tool was divided into 2 distinct parts:

#### Part I

Socio-demographic information.

#### Part II

Knowledge regarding safe abortion among married reproductive age women of Vyas municipality-9, Tanahun. It consists of 14 questions that helped to assess the level of knowledge on safe abortion. Level of knowledge was classified into 2 categories. Adequate knowledge (more than the mean value) and inadequate knowledge (equal to or less than the mean value).

### Technique of Data Collection

Permission was taken from concerned authority. Informed consent was taken from participants and objectives of the study was explained to the participants. Data was collected through self-developed semi structured questionnaire via face-to-face interview technique.

The time taken to interview each participant was about 20-25 minutes.

### Pre-Testing Tools

The pretesting was done among 10% (12) of the sample size in a similar setting. It was conducted among married women of reproductive age group of Vyas municipality-7, Tanahun, and necessary correction was done as needed.

Validity and Reliability of Instruments

#### Validity

The content validity was done through an extensive literature review, consulting research guide and related teachers.

#### Data Collection Procedure

Before data collection, a formal approval letter was obtained from the concerned authority of PUSHS.

A formal letter from PUSHS was submitted to the Vyas Municipality office.

After accepting permission from the concerned authority of the Vyas Municipality office, the following step were taken up:

Step 1: Self introduction was given.

Step 2: Explanation of the objectives of the study to the respondents.

Step 3: Written consent was taken from the respondents.

Step 4: Participants privacy and convenience were ensured with proper confidentiality.

Step 5: Data were collected by face-to-face semi-structured interview technique.

Step 6: Nepali language was used.

Step 7: 15-20 minutes were taken for each respondent.

Step 8: Data were collected within 2 weeks.

### Data Analysis Procedure

1. Collected data was checked for completeness and accuracy then categorized.
2. The data was arranged, coded and kept in order for editing and entered into SPSS 16.0 version.
3. The data was analyzed through descriptive statistics i.e. frequency, percentage, mean, standard deviation, and chi-square test was used to find out the association between level of knowledge, and selected socio-demographic variables. Interpretation of data was presented through tables.

### Ethical Consideration

Formal permission was obtained from the higher authority of PUSHS. Formal permission was obtained from the municipality. The purpose of the study was explained to all respondents. Informed written consent was taken from respondents before data collection. Anonymity and confidentiality were maintained by not disclosing the information provided by respondents. The respondents were ensured that the information was used for research purposes only.

## Results

### Socio-demographic characteristics of the respondents

Table 1 shows that among 118 respondents, 28% was age between 25-29 years, with a mean age ± SD of 30.81±6.704. Almost all (92.4%) of the respondents follow the Hindu religion, and more than one third (39.8%) of the respondents were from Janajati ethnicity. Almost all (94.9%) of the respondents can read and write and nearly less than half (49.1%) of the respondents have secondary level education. Two-thirds (61%) of the respondents were homemakers, and more than half (59.3%) of the respondents belonged to a joint family. Two thirds (61%) of the respondent’s families earn less than 35000 monthly.

**Table 1:**
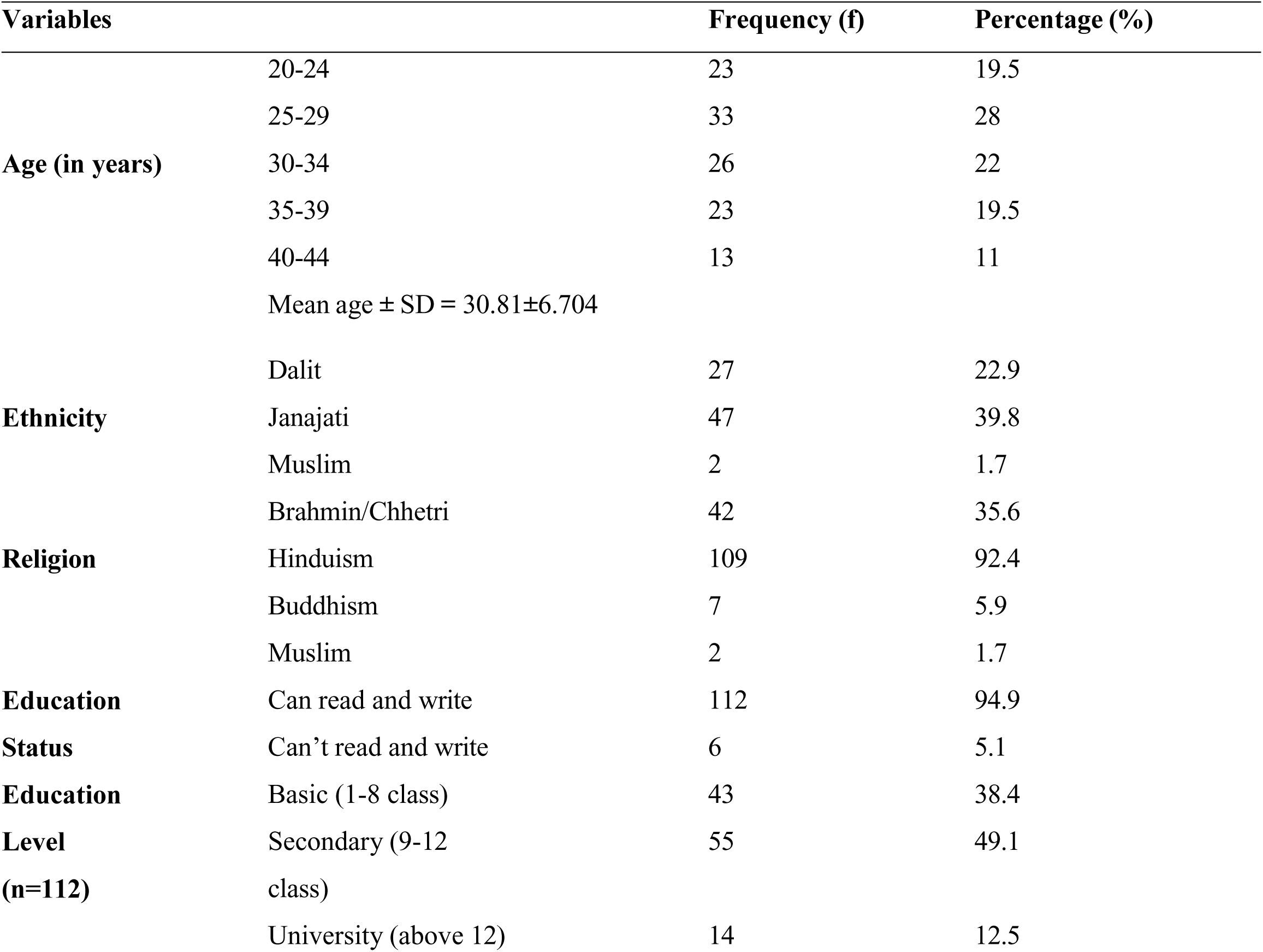
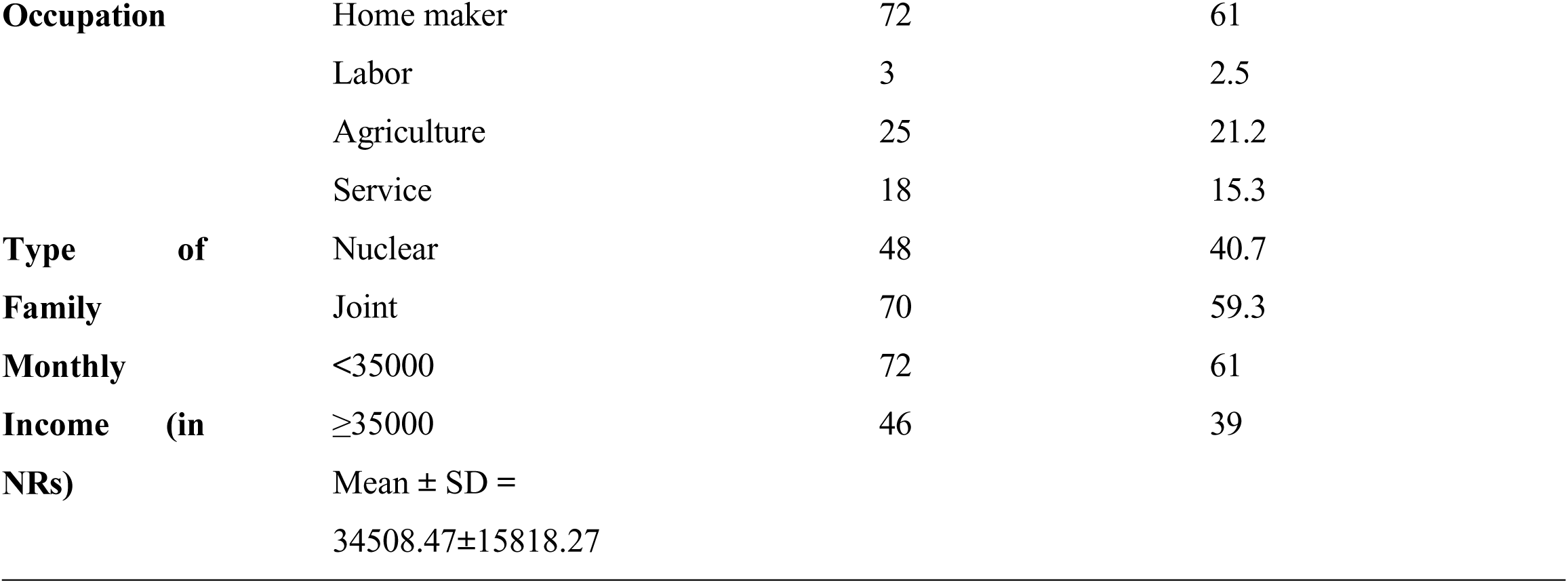
Socio-demographic characteristics of the respondents. (n=118)

### Source of Information of the Respondents

Table 2 shows that all (100%) of the respondents heard about abortion, and the majority (87.3%) of the respondent’s sources of information about abortion were from friends and relatives. Less than one third (25.4%) of the respondents had a history of abortion, and more than two third (66.7%) of the respondent the main cause was unwanted pregnancy.

**Table 2:**
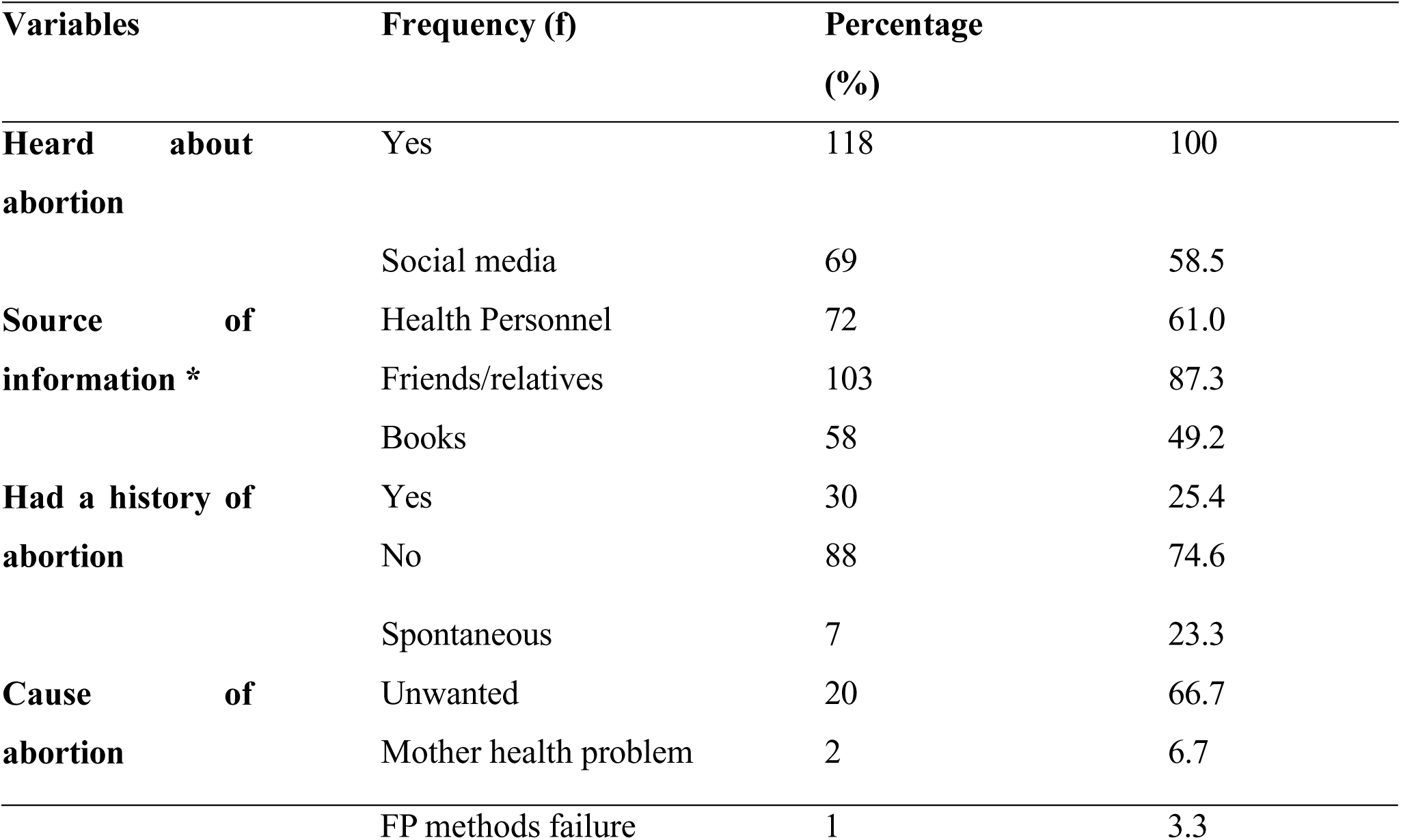
Source of Information of the Respondents.

### Knowledge regarding safe abortion among the respondents

Table 3 presents the respondents’ knowledge regarding safe abortion among 118 respondents. More than half (53.4%) of the respondents correctly identified the definition of abortion and majority (86.4%) of the respondents heard about type of abortion, almost all (97.5%) of the respondents correctly identified trauma or injury cause of abortion, majority (86.4%) of the respondents correctly answered the methods of abortion. Almost all (99.2%) of the respondents understand safe abortion, majority (86.4%) of the respondents know the safe time frame for a safe abortion.

**Table 3:**
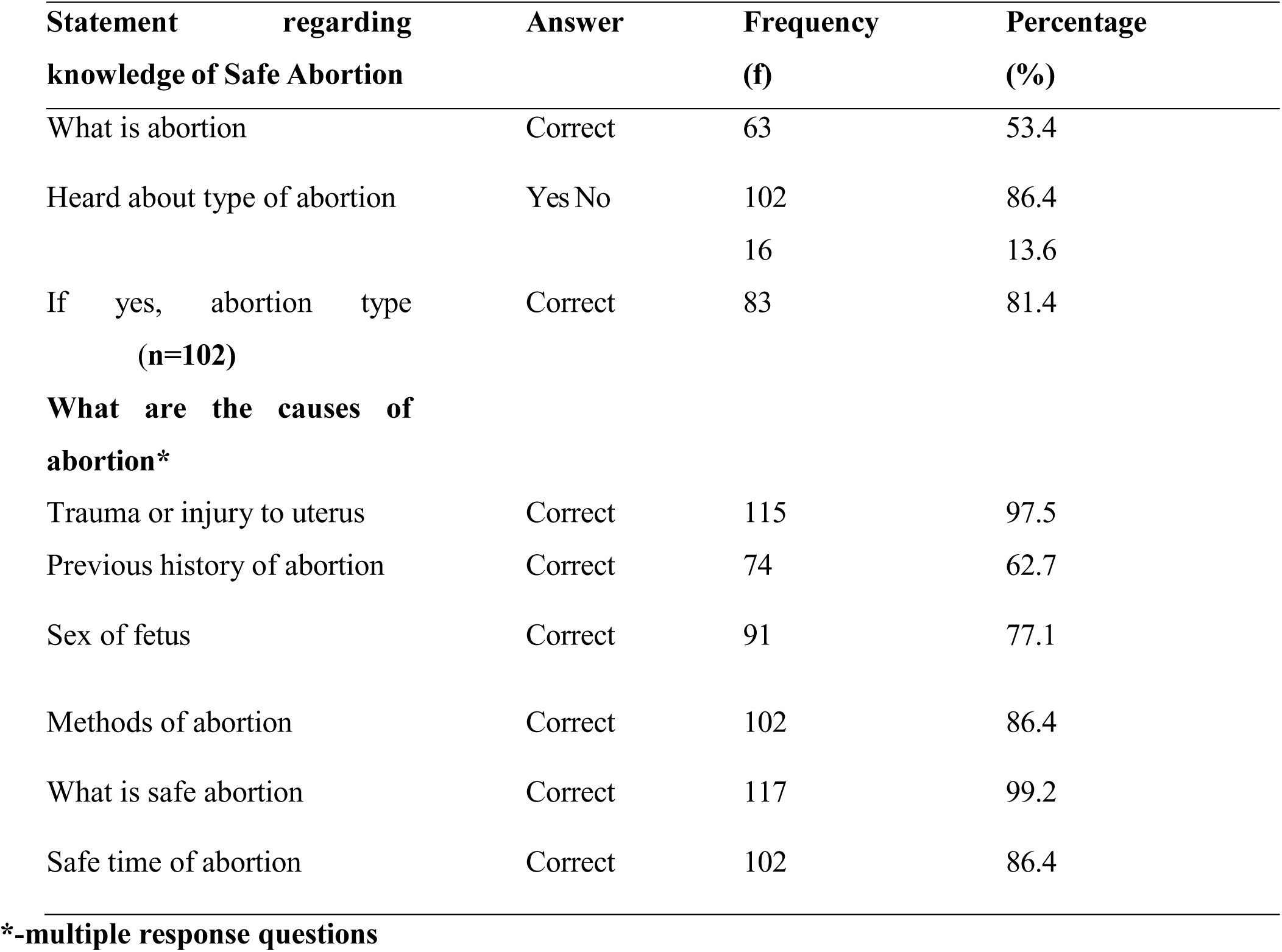
Knowledge regarding safe abortion among the respondents.

### Knowledge regarding safe abortion among the respondents

Table 4 shows that almost all (93.2%) of the respondents know about government services. All respondents are aware of unsafe abortion, where the majority (86.4%) of the respondents correctly answered about unsafe abortion. All respondents are aware of complications of vaginal bleeding; almost all (94.9%) of the respondents know about the law, and only half are aware of the specific legal conditions for abortion in Nepal.

**Table 4:**
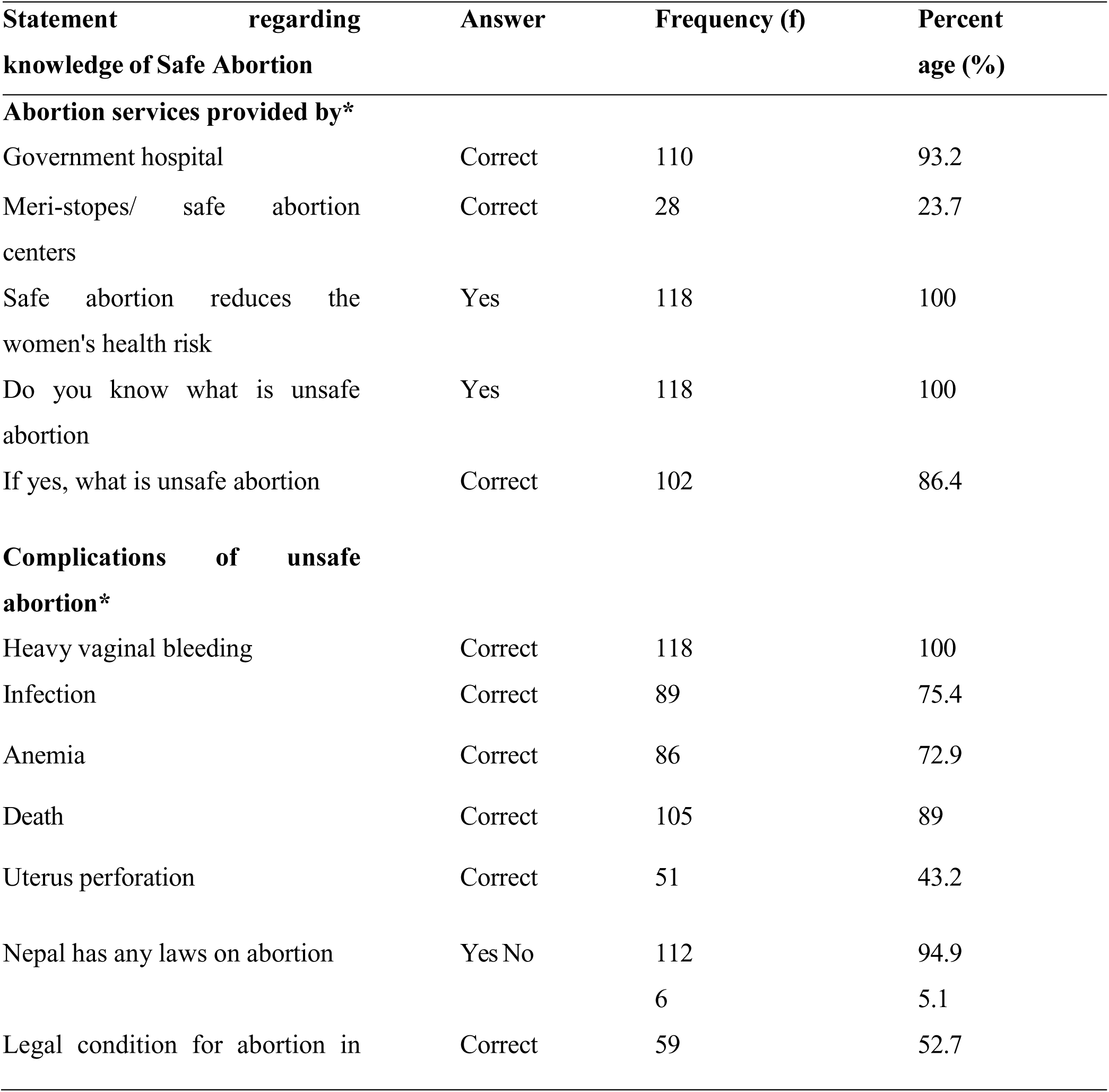
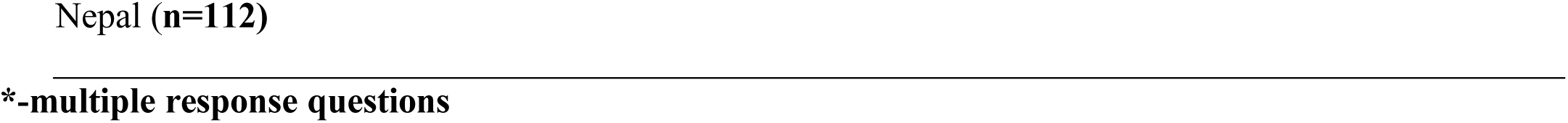
Knowledge regarding safe abortion among the respondents.

| Statement regarding knowledge of Safe Abortion | Answer | Frequency (f) | Percent age (%) |
| --- | --- | --- | --- |
| **Abortion services provided by\*** |  |  |  |
| Government hospital | Correct | 110 | 93.2 |
| Meri-stopes/ safe abortion centers | Correct | 28 | 23.7 |
| Safe abortion reduces the women's health risk | Yes | 118 | 100 |
| Do you know what is unsafe abortion | Yes | 118 | 100 |
| If yes, what is unsafe abortion | Correct | 102 | 86.4 |
| **Complications of unsafe abortion\*** |  |  |  |
| Heavy vaginal bleeding | Correct | 118 | 100 |
| Infection | Correct | 89 | 75.4 |
| Anemia | Correct | 86 | 72.9 |
| Death | Correct | 105 | 89 |
| Uterus perforation | Correct | 51 | 43.2 |
| Nepal has any laws on abortion | Yes No | 112 | 94.9 |
|  |  | 6 | 5.1 |
| Legal condition for abortion in | Correct | 59 | 52.7 |

**\*-multiple response questions**

### Level of knowledge regarding safe abortion among the respondents

Table 5 shows the overall knowledge of respondents regarding safe abortion. Among the total respondents, nearly two third (57.6%) of the respondents had adequate knowledge and less than half (42.4%) of the respondents had inadequate knowledge regarding safe abortion.

**Table 5:** Level of knowledge regarding safe abortion among the respondents.

| Level of knowledge | Frequency (n) | Percentage (%) |
| --- | --- | --- |
| Adequate | 68 | 57.6 |
| Inadequate | 50 | 42.4 |

### Association of level of knowledge regarding safe abortion with selected demographic variables

Test statistics: Pearson chi-square and linear-by-linear value, *p-value significance at < 0.05 Table 6 shows that the level of knowledge regarding safe abortion is significantly associated with education status (p=0.038) of the respondents, with p-value <0.05.

**Table 6.**
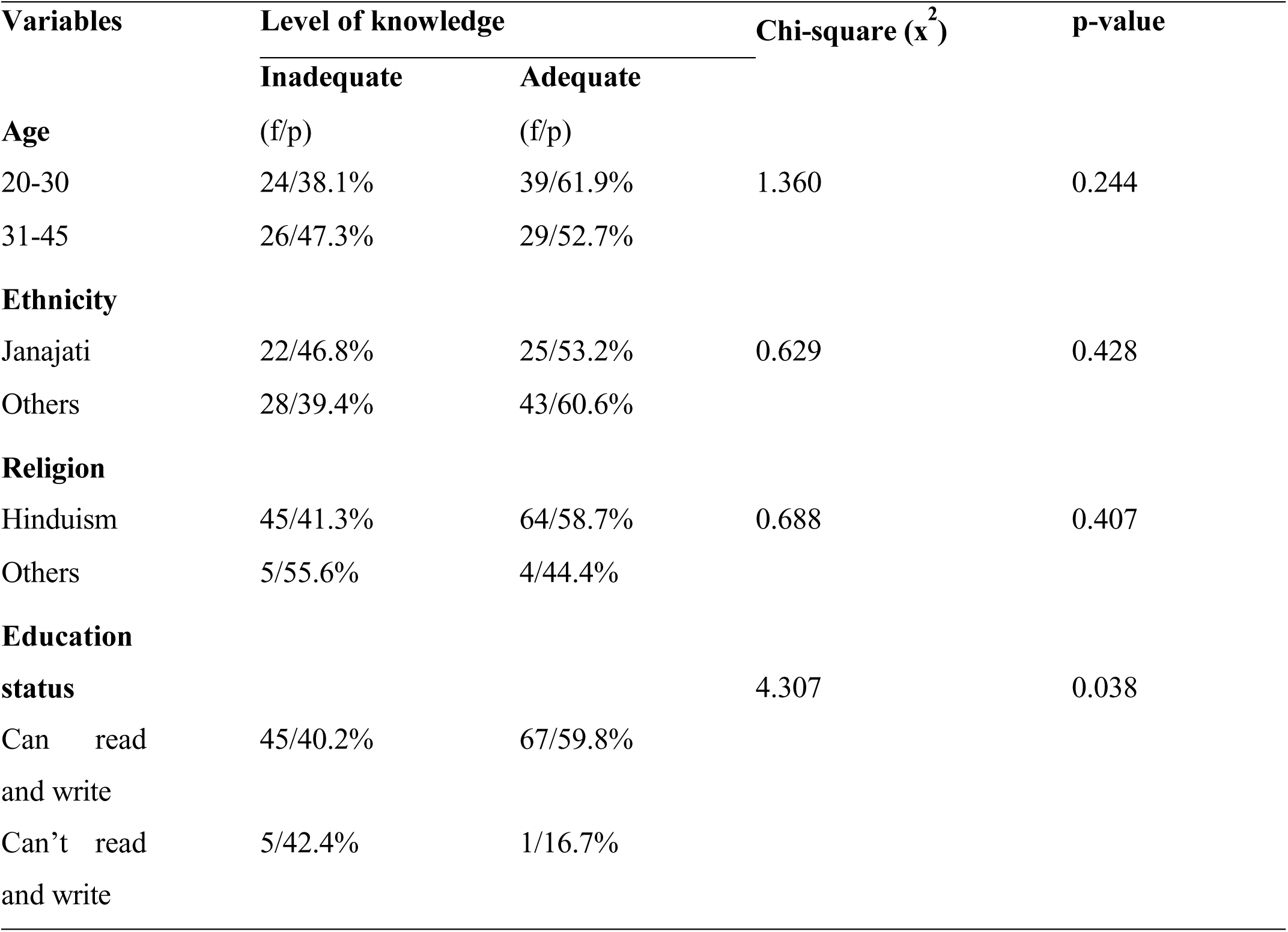
Association of level of knowledge regarding safe abortion with selected demographic variables. (n=118)

Test statistics: Pearson chi-square and linear-by-linear value, *p-value significance at < 0.05 Table 7 shows that level of knowledge regarding safe abortion is significantly associated with and education level (0.002) of the respondents. It shows that the knowledge increases with education.

**Table 7:**
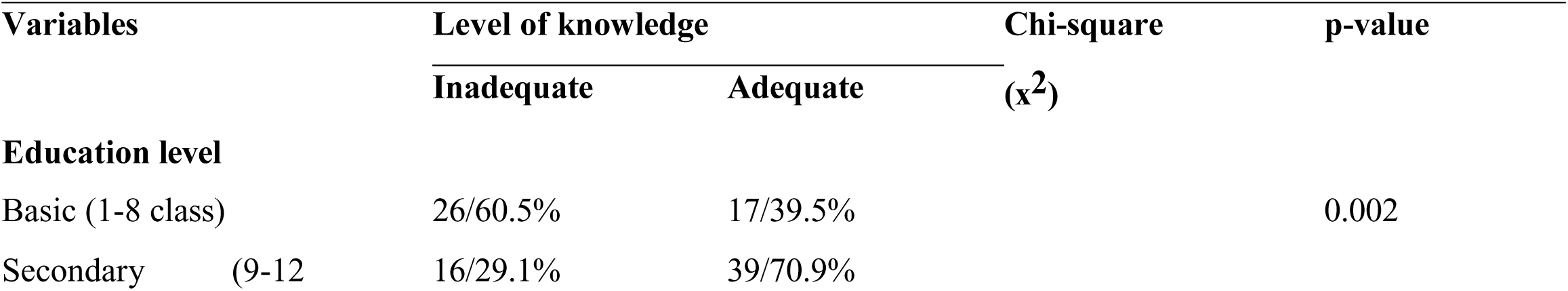
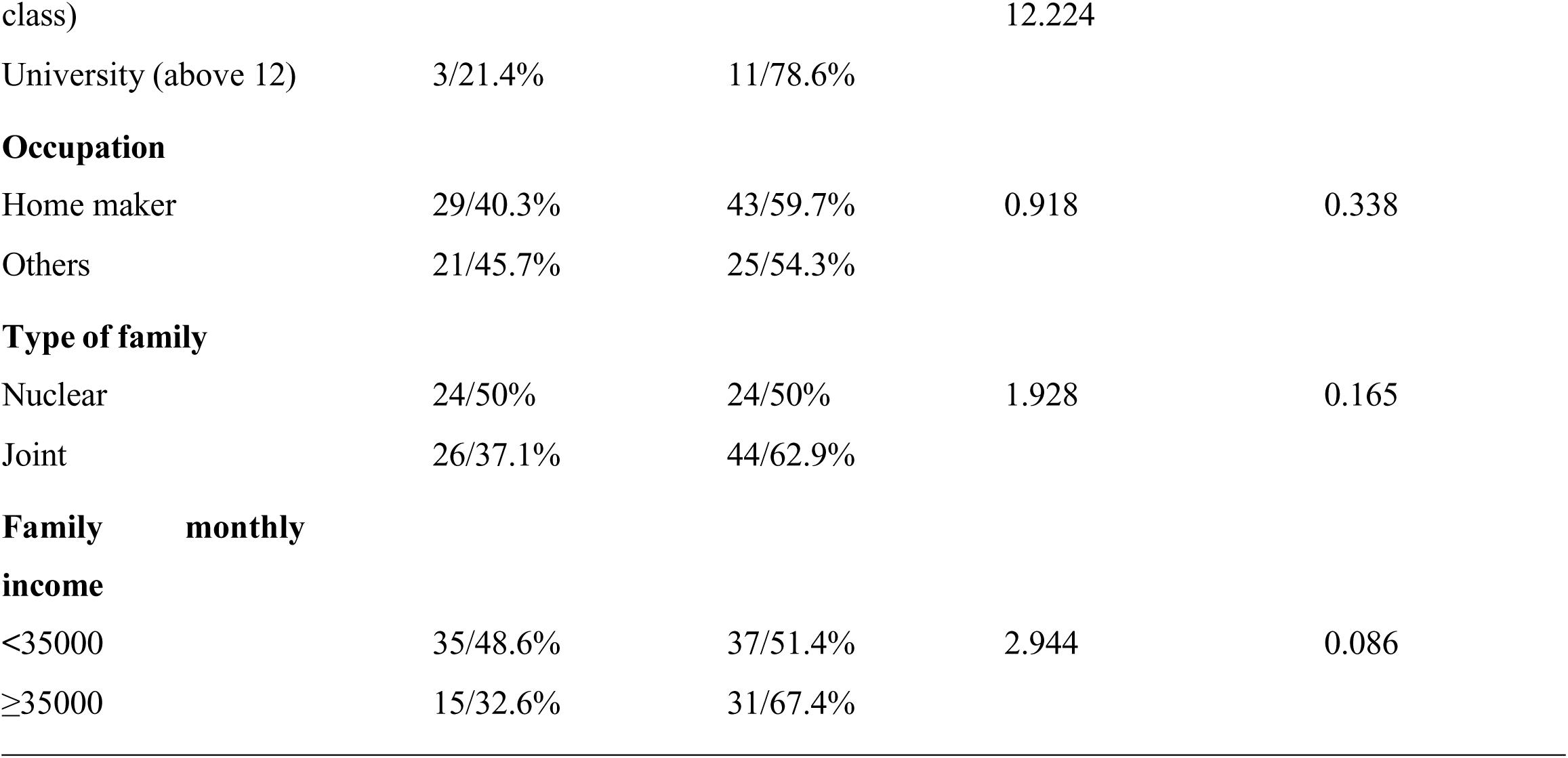
Association of level of knowledge regarding safe abortion with selected demographic variables.

## Discussion

The aim of the study was to assess the level of knowledge regarding safe abortion among married women of reproductive age in Vyas-9, Tanahun. A descriptive cross-sectional study design was conducted among 118 women to assess the level of knowledge regarding safe abortion and to find out the association between levels of knowledge on safe abortion with selected demographic variables. The study population was reproductive age women between 20-45 years and non-probability sampling technique was used. Data was collected by using self-constructed semi structured questionnaire via face-to-face interview technique.

In this study, 11% of respondents were between age group of 40-45 years. This finding was supported by the study which was conducted at Lekhnath, Kaski, which revealed that 6% of respondents were from 40-44 years (Acharya A *et al.,* 2017). Both studies show that older women were less represented in the study. This similarity is due to the fact that women closer to menopause may be less involved in health discussion or abortion decisions, leading to less participation.

The present study shows that less than half (39.8%) of the respondents from Janajati ethnic group, and 5.9% of the respondents were from Buddhism religion. Among the total respondents, 12.5% were from higher education level. This finding was supported by the study done in slum area of Kathmandu, which shows that 43.1% of the respondents belongs to Janajati ethnic group, 4.7% of the respondents were from Buddhism religion and minority of respondents 6.3% were from higher education level (Manandhar N *et al.,* 2021). The similarity results between two studies reflect the same diversity in Nepal population, and semi-urban and slum areas often lead to limited access to higher education.

This current study reveals that majority (86.4%) of the respondents answered correctly on safe gestational period to undergo induced abortion. This finding was contradicted by the study done in Heart, Afganistan, which shows that more than half (56.6%) of the respondents were correctly answered on safe time for abortion (Neyazi A *et al.,* 2021). The lower level of knowledge on safe gestational period to undergo induced abortion in Afghanistan study could be attributed to high illiteracy rate among respondents, as more than half of them were unable to read or write.

This current study illustrates that almost all (93.2%) of the respondents answered safe abortion service was provided by government hospital. More than two third (65.8%) of the respondents knew the infection was the complication of unsafe abortion. More than half (52.7%) of the respondents correctly answered the legal condition for abortion in Nepal. This finding was supported by the study done in Pokhara Metropolitan city, which shows the almost all (93.3%) of the respondents knew the government hospital was the safe abortion site. Similarly, 75.4% of the respondent answered infection was the one of the complications of unsafe abortion. Nearly two third (59.6%) of the respondents knew the legal condition for safe abortion (Thapaliya R *et al.,* 2024). The similarity in findings may be attributed to consistent reproductive health messaging across Nepal, where infection is widely acknowledged as a primary complication of unsafe abortion.

The present study reveals that nearly two third (57.6%) of the respondents had adequate knowledge on safe abortion, while less than half (42.4%) of the respondents had inadequate knowledge. This study finding was supported by the study done in Herat, Afganistan on Abortion, which shows that 56.6% of the total respondents had good level of knowledge on safe abortion and 43.4% of the respondents had poor knowledge (Neyazi A *et al.,* 2021). The both studies found that more than half of the respondents had good knowledge. Despite different countries, this indicating the common global health trends, education and exposure to health facilities influence the knowledge on safe abortion. The current study reveals, a significant association was found between knowledge levels and education status and level (p<0.05) and not significant with other demographic variables age, religion, ethnicity and occupation. These findings are consistent with finding of a study done in Afganistan. In their study also found the most of the demographic variables were insignificant with the level of knowledge (Saeed S *et al.,* 2022). Both studies illustrate that education is universally linked to higher health literacy, higher levels of education are consistently associated with improved health awareness.

## Conclusion

The present study concludes that nearly two third of the respondents have adequate knowledge regarding safe abortion. Educational status significantly influenced their level of knowledge. Thus, if we could provide correct knowledge, education and enhancing awareness to married women of reproductive aged group, it can help to reduce unsafe abortion and improve reproductive health outcomes.

## Implication

1. The findings of the study can be implemented in the health care centers, education, administration and research area.
2. The findings of the study help to develop plan and policies about safe abortion.

## Data Availability

All data produced (questionnaire) are available online at figshare

https://figshare.com/authors/Dipesh_Thapa/23756241

